# Association Between In-Hospital Antibiotic Use and Long-Term Outcomes in Critically Ill Patients

**DOI:** 10.1101/2025.03.24.25324548

**Authors:** Parker Burrows, Ruth-Ann Brown, Abigail Samuelsen, Anthony S Bonavia

**Author notes:** Corresponding author:* Anthony Bonavia, MD FCCP. Funding:* This study was funded by the National Institute of General Medical Sciences, grants # K08GM138825 and R35GM150695 (ASB). Data Availability:* Data are available from the authors on reasonable request. Ethical Approval* was provided by The Penn State College of Medicine Institutional Review Board of the Human Studies Protection Office on July 30, 2020 (ref# 15328). All methods were carried out in accordance with relevant institutional guidelines and regulations.

## Abstract

**Objective:** To assess whether antibiotic duration (AD) and one-year antibiotic-free days (AFD) are associated with key in-hospital and post-discharge outcomes among critically ill adults.

**Design:** Retrospective observational study.

**Setting:** Quaternary care academic medical center in the United States.

**Patients:** A total of 126 critically ill adults, mean age 68.1 years (±15.6), 51.6% male, median APACHE II score of 20.5 (IQR 15–25); 71.4% met sepsis criteria.

**Methods:** Patient demographics, clinical characteristics, antibiotic use, and outcomes were collected over one year. Secondary infection was defined as ≥3 consecutive antibiotic days within a year following the index sepsis admission. Multivariate analyses adjusted for age, APACHE II score, gender, and glucocorticosteroid dose.

**Results:** Within 30 days, longer AD correlated with increased hospital stay (p<0.001) with each additional day of antibiotics associated with 0.37 – 0.39 extra days of hospitalization in univariate and multivariate analyses, respectively. In septic patients specifically, AFD significantly correlated with hospital length-of-stay in both univariate (p=0.023) and multivariate analyses (p=0.002), with no impact from infection type on AD or AFD. Fewer AFD correlated with higher secondary bacteremia rates in unadjusted analysis (p=0.023 overall), but this effect was not significant after multivariable adjustment. Neither AD nor AFD predicted one-year mortality or readmission.

**Conclusions:** Extended antibiotic duration in critically ill patients prolonged hospital stays without providing mortality or readmission benefits. These findings underscore the importance of robust antibiotic stewardship, where shorter, targeted regimens can reduce unintended complications and improve overall outcomes.

## Introduction

Sepsis remains a major cause of morbidity and mortality in critically ill populations worldwide, yet its diagnosis is inherently clinical.^1^ Current consensus guidelines rely on the Sequential Organ Failure Assessment (SOFA) score^2^ to operationalize organ dysfunction; however, no biomarkers definitively diagnose sepsis as of yet. The inherent complexity of sepsis pathophysiology and the absence of a gold-standard diagnostic biomarker create both diagnostic uncertainty and therapeutic urgency.

Given the documented survival benefit of early antibiotic initiation,^3–5^ clinicians often err on the side of rapidly administering broad-spectrum antibiotics when sepsis is suspected, even in the absence of positive microbial culture results. This approach is clinically justified because culture results can take days to mature, and delaying antibiotics beyond the initial window can substantially increase mortality.^6,7^ Under these circumstances, starting antibiotics preemptively is particularly common in critically ill and high-risk patients, in whom even a short delay might prove detrimental. Nevertheless, the challenge arises when empirically administered antibiotics are subsequently continued longer than clinically warranted, leading to inappropriately prolonged antibiotic courses that do not necessarily align with the patient’s evolving clinical picture or definitive culture data.

One of the most concerning consequences of extended antibiotic therapy, especially when not guided by confirmed microbial etiology, is its potential to disrupt the host microbiome. Antibiotics can alter both whole-body and organ-specific microbial communities, affecting gut flora and other mucosal surfaces in ways that may predispose patients to immune dysregulation and secondary infections.^8^ Although antibiotic regimens are indispensable for treating bacterial infections, an accumulating body of literature has demonstrated that the detrimental effects of antibiotic overexposure may offset potential therapeutic benefits, especially when continued for longer durations than indicated. ^9–11^

Given these complexities, the goal of the present study was to perform a descriptive analysis of the long-term (up to one year) outcomes associated with different antibiotic duration (AD) courses in critically ill patients. We hypothesized that prolonged antibiotic exposure would increase the risk of secondary infections and hospital resource utilization, without necessarily improving mortality. Additionally, we sought to define “infection” in a manner that more reliably distinguishes empiric therapy from continued therapy in response to a probable or confirmed infection. Specifically, our criterion for secondary infection was the receipt of at least three consecutive days of antibiotics. This threshold reflects our assumption that a regimen extended to or beyond three days implies a strong clinical suspicion of infection rather than a brief “cover-until-ruled-out” approach.^12,13^ We do not presume that this cutoff defines an optimal AD for any given pathogen; rather, it serves as a practical tool for discerning short-term empiric treatment from courses intended for active infection management.

By detailing one-year outcomes—such as secondary infections, bacteremia, mortality, and readmission—this study addresses a gap in the current antibiotic stewardship literature, where much of the focus has been placed on shorter-term results. The findings may further inform best practices surrounding antibiotic prescriptions in the intensive care unit, encouraging more nuanced decisions that weigh the benefits of early empiric therapy against the hazards of prolonged and potentially unnecessary antibiotic exposure.

## Results

### Cohort Characteristics

A total of 126 critically ill adults were included, of whom 90 (71.4%) met sepsis criteria and 36 (28.6%) were classified as non-septic. **Table 1** presents a detailed comparison of demographic and clinical features between these two subgroups. The mean age was 68.1 years in both groups, and there was no significant difference in sex distribution (47.7% female in the septic group vs. 50.0% female in the non-septic group, p=0.82). Race, body mass index, and severity-of-illness measures (Acute Physiology and Chronic Health Evaluation II [APACHE II] and SOFA scores) were also comparable. Leukocyte counts at admission were notably higher among septic patients (median 16.7 ×10^3^/µL [IQR 12.2–23.5] vs. 10.7 ×10^3^/µL [IQR 7.6–18.3], p<0.001), and lactic acid levels were mildly elevated in the sepsis cohort (median 3.0 mg/dL [IQR 1.8–4.9] vs. 2.6 mg/dL [IQR 1.6–4.5], p=0.04). Although septic patients more frequently required vasopressors at admission, the difference did not reach statistical significance (48.9% vs. 33.3%, p=0.11).

**Table 1:**
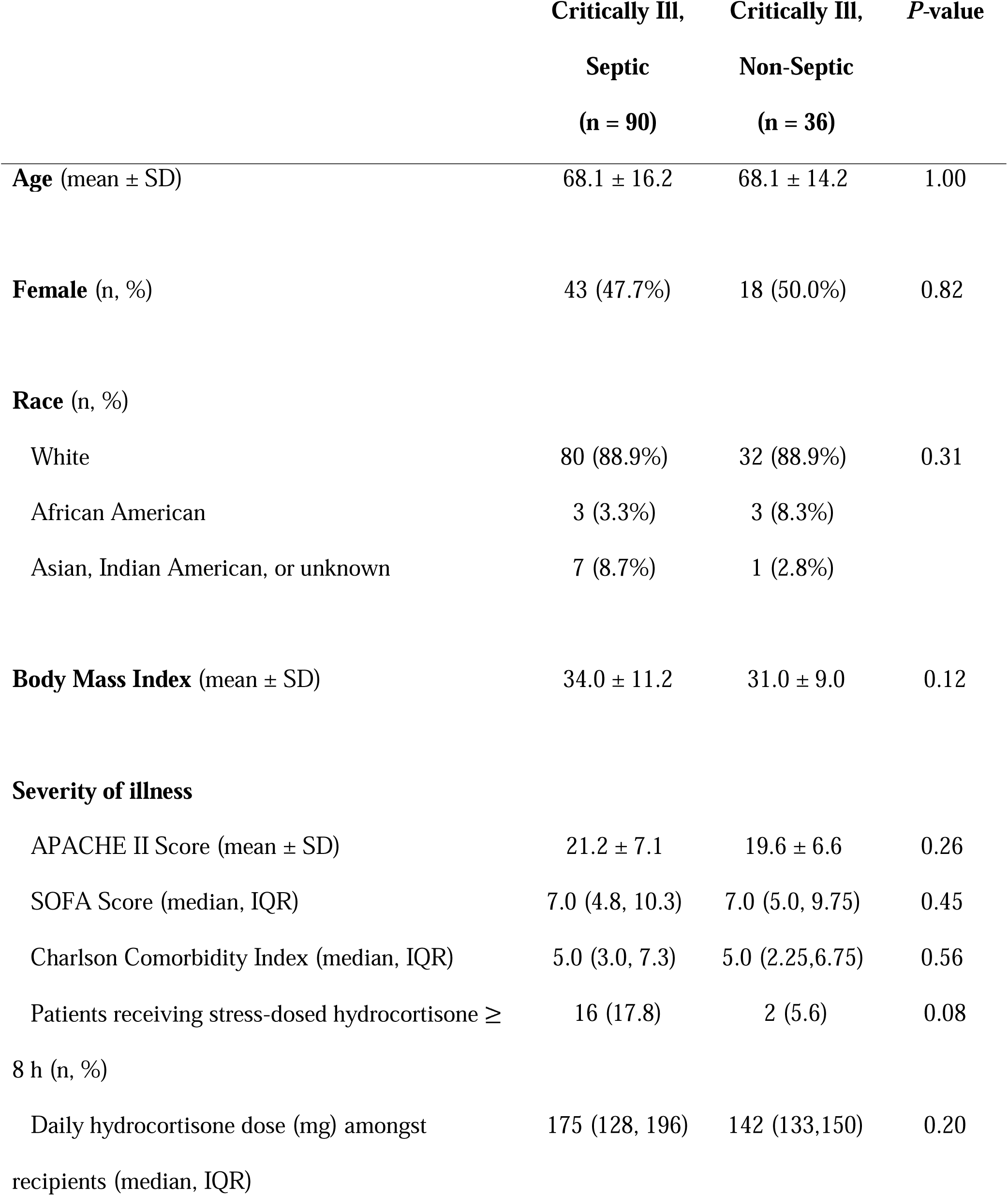
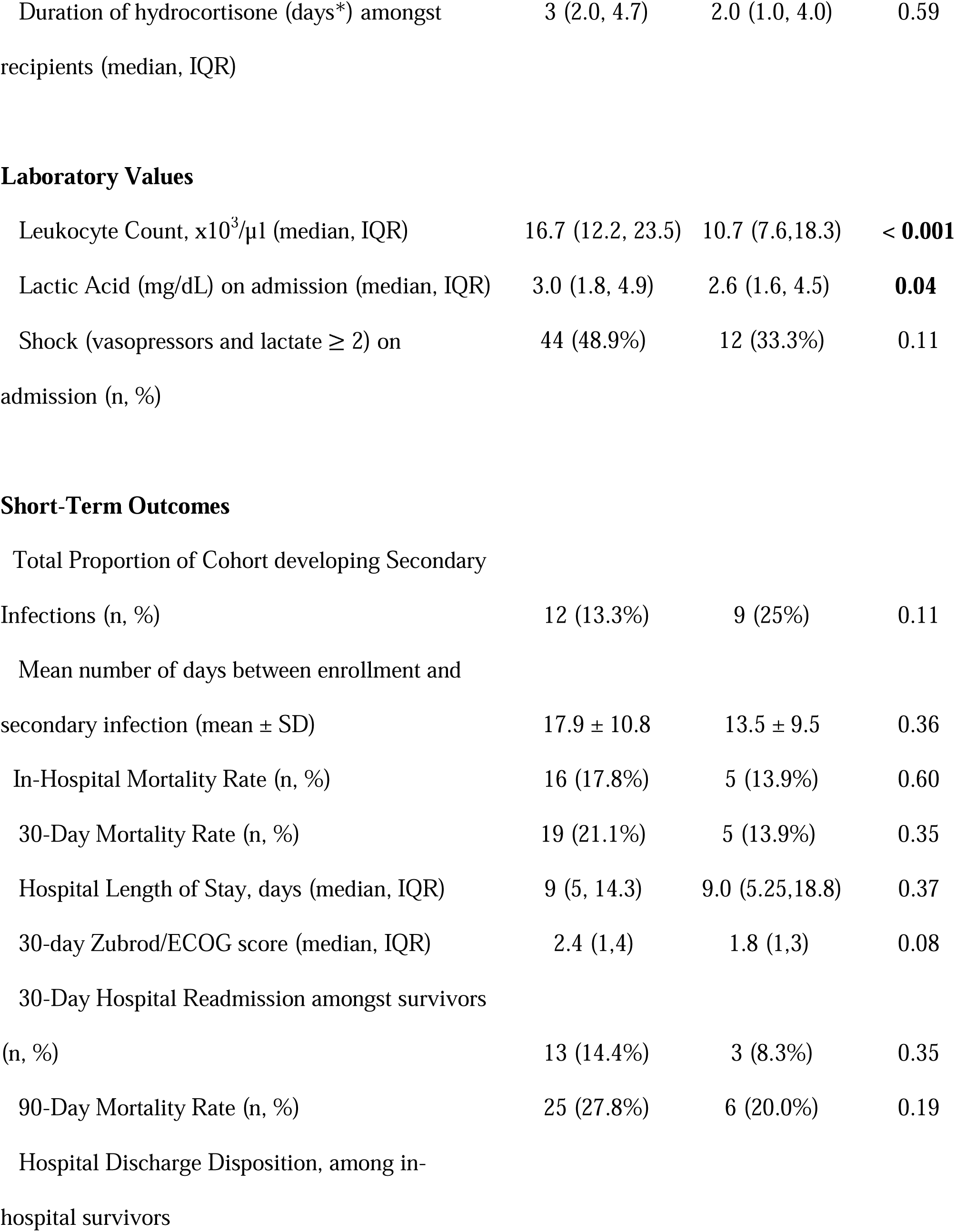
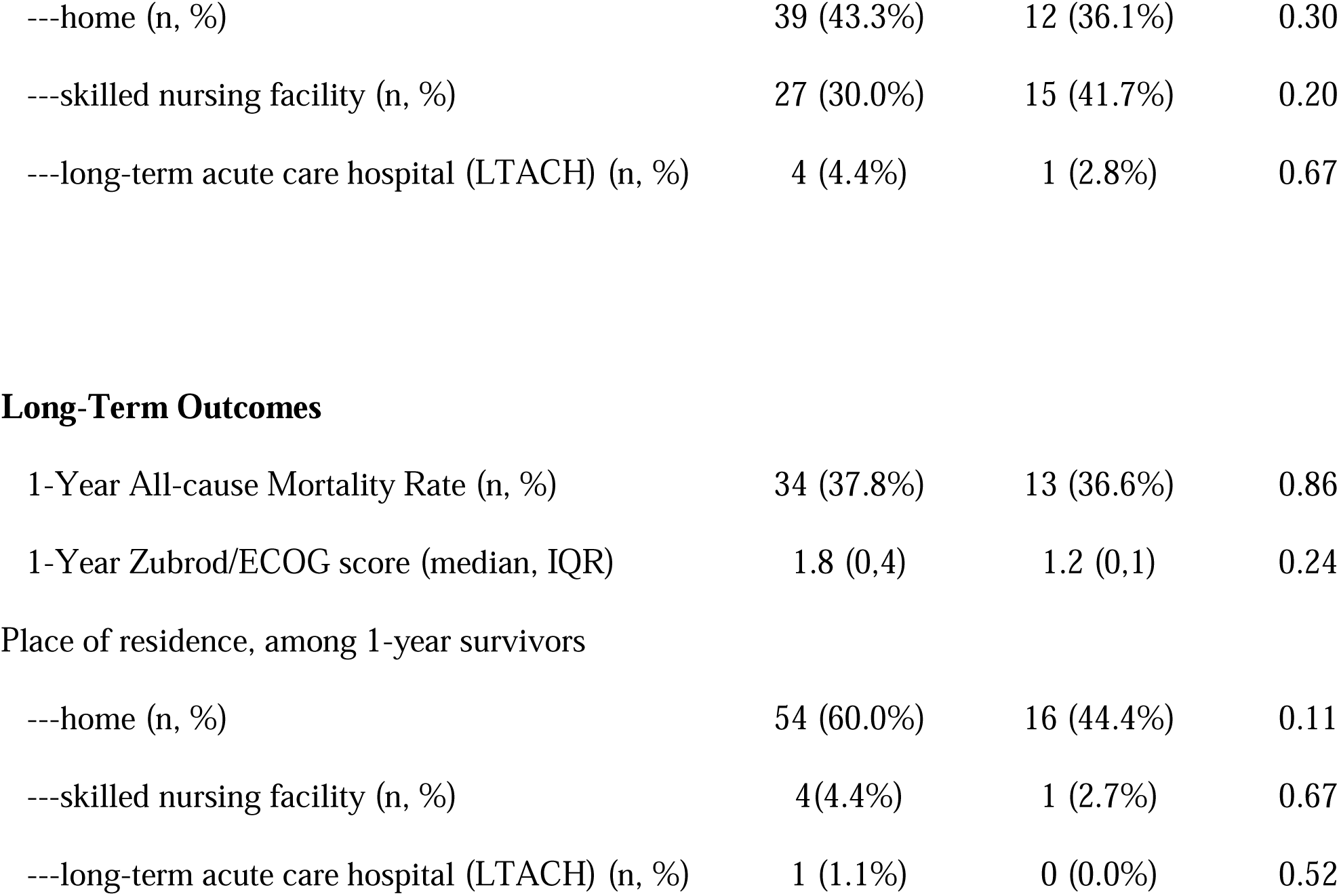
Patient Demographics and Outcomes.

|  | **Critically Ill,**<br>**Septic**<br>**(n = 90)** | **Critically Ill,**<br>**Non-Septic**<br>**(n = 36)** | ***P*-value** |
| --- | --- | --- | --- |
| **Age** (mean ± SD) | 68.1 ± 16.2 | 68.1 ± 14.2 | 1.00 |
| **Female** (n, %) | 43 (47.7%) | 18 (50.0%) | 0.82 |
| **Race** (n, %) |  |  |  |
| White | 80 (88.9%) | 32 (88.9%) | 0.31 |
| African American | 3 (3.3%) | 3 (8.3%) |  |
| Asian, Indian American, or unknown | 7 (8.7%) | 1 (2.8%) |  |
| **Body Mass Index** (mean ± SD) | 34.0 ± 11.2 | 31.0 ± 9.0 | 0.12 |
| **Severity of illness** |  |  |  |
| APACHE II Score (mean ± SD) | 21.2 ± 7.1 | 19.6 ± 6.6 | 0.26 |
| SOFA Score (median, IQR) | 7.0 (4.8, 10.3) | 7.0 (5.0, 9.75) | 0.45 |
| Charlson Comorbidity Index (median, IQR) | 5.0 (3.0, 7.3) | 5.0 (2.25,6.75) | 0.56 |
| Patients receiving stress-dosed hydrocortisone ≥ 8 h (n, %) | 16 (17.8) | 2 (5.6) | 0.08 |
| Daily hydrocortisone dose (mg) amongst recipients (median, IQR) | 175 (128, 196) | 142 (133,150) | 0.20 |
| Duration of hydrocortisone (days*) amongst recipients (median, IQR) | 3 (2.0, 4.7) | 2.0 (1.0, 4.0) | 0.59 |
| Laboratory Values |  |  |  |
| Leukocyte Count, $\times 10^3/\mu\text{l}$ (median, IQR) | 16.7 (12.2, 23.5) | 10.7 (7.6, 18.3) | < **0.001** |
| Lactic Acid (mg/dL) on admission (median, IQR) | 3.0 (1.8, 4.9) | 2.6 (1.6, 4.5) | **0.04** |
| Shock (vasopressors and lactate $\geq 2$) on admission (n, %) | 44 (48.9%) | 12 (33.3%) | 0.11 |
| Total Proportion of Cohort developing Secondary Infections (n, %) | 12 (13.3%) | 9 (25%) | 0.11 |
| Mean number of days between enrollment and secondary infection (mean ± SD) | 17.9 ± 10.8 | 13.5 ± 9.5 | 0.36 |
| In-Hospital Mortality Rate (n, %) | 16 (17.8%) | 5 (13.9%) | 0.60 |
| 30-Day Mortality Rate (n, %) | 19 (21.1%) | 5 (13.9%) | 0.35 |
| Hospital Length of Stay, days (median, IQR) | 9 (5, 14.3) | 9.0 (5.25, 18.8) | 0.37 |
| 30-day Zubrod/ECOG score (median, IQR) | 2.4 (1, 4) | 1.8 (1, 3) | 0.08 |
| 30-Day Hospital Readmission amongst survivors (n, %) | 13 (14.4%) | 3 (8.3%) | 0.35 |
| 90-Day Mortality Rate (n, %) | 25 (27.8%) | 6 (20.0%) | 0.19 |
| Hospital Discharge Disposition, among in-hospital survivors |  |  |  |
| ---home (n, %) | 39 (43.3%) | 12 (36.1%) | 0.30 |
| ---skilled nursing facility (n, %) | 27 (30.0%) | 15 (41.7%) | 0.20 |
| ---long-term acute care hospital (LTACH) (n, %) | 4 (4.4%) | 1 (2.8%) | 0.67 |
| **Long-Term Outcomes** |  |  |  |
| 1-Year All-cause Mortality Rate (n, %) | 34 (37.8%) | 13 (36.6%) | 0.86 |
| 1-Year Zubrod/ECOG score (median, IQR) | 1.8 (0,4) | 1.2 (0,1) | 0.24 |
| **Place of residence, among 1-year survivors** |  |  |  |
| ---home (n, %) | 54 (60.0%) | 16 (44.4%) | 0.11 |
| ---skilled nursing facility (n, %) | 4(4.4%) | 1 (2.7%) | 0.67 |
| ---long-term acute care hospital (LTACH) (n, %) | 1 (1.1%) | 0 (0.0%) | 0.52 |

### Infectious Etiology

Primary infections among the septic cohort included gram-positive (30.0%), gram-negative (33.3%), mixed (16.7%), and fungal or viral etiologies (7.7%). **Table 2** summarizes infectious etiologies at both the index hospitalization and for secondary infections, delineating whether pathogens were gram-positive, gram-negative, fungal, viral, or clinically diagnosed without microbial confirmation. **Supplementary Table 1** enumerates the infectious etiologies and corresponding sources of infection. Overall, pneumonia and bacteremia were among the most common initial presentations, while secondary infections varied from respiratory to urinary and skin or soft-tissue sites.

**Table 2:** Infectious Etiology and Sources.

|  | **Critically**<br>**Ill, Septic** | **Critically**<br>**Ill, Non-**<br>**Septic** | ***P*-value** |
| --- | --- | --- | --- |
| **Etiology of Secondary infections < 30 days** | N= 14 | N= 9 | 0.48 |
| Gram-positive | 2 (14.3%) | 3 (33.3%) |  |
| Gram-negative | 2 (14.3%) | 2 (22.2%) |  |
| Fungal | 2 (14.3%) | 0 (0.0%) |  |
| Viral | 1 (7.1%) | 1 (11.1%) |  |
| Mixed | 4 (28.6%) | 3 (33.3%) |  |
| Clinical Diagnosis of Infection only | 3 (21.4%) | 0 (0.0%) |  |
| **Etiology of Secondary infections 31 – 365 days** | N=32 | N=7 | 0.40 |
| Gram-positive | 11 (34.4%) | 2 (28.6%) |  |
| Gram-negative | 7 (21.9%) | 4 (57.1%) |  |
| Fungal | 0 (0.0%) | 0 (0.0%) |  |
| Viral | 2 (6.3%) | 0 (0.0%) |  |
| Mixed | 3 (9.4%) | 1 (14.3%) |  |
| Clinical Diagnosis of Infection only | 9 (20.6%) | 0 (0.0%) |  |
| **Source of Secondary infections < 30 days** | N= 11 | N= 10 | 0.70 |
| Pneumonia | 3 (27.3%) | 4 (40.0%) |  |
| Abdominal Infection | 1 (9.1%) | 1 (10.0%) |  |
| Urinary tract infection | 1 (9.1%) | 2 (20.0%) |  |
| Skin or Soft Tissue Infection | 6 (54.5%) | 3 (30.0%) |  |
| Other | 0 (0.0%) | 0 (0.0%) |  |
| **Source of Secondary infections 31 – 365 days** | **N=27** | **N= 6** | **0.37** |
| Pneumonia | 4 (14.8%) | 2 (33.3%) |  |
| Abdominal Infection | 0 (0.0%) | 0 (0.0%) |  |
| Urinary tract infection | 12 (44.4%) | 1 (16.7%) |  |
| Skin or Soft Tissue Infection | 11 (40.7%) | 3 (50.0%) |  |
| Other | 0 (0.0%) | 0 (0.0%) |  |

### Short-Term Outcomes

Short-term clinical endpoints, including secondary infections, in-hospital mortality, and 30-day mortality, are summarized in **Table 1**. Within 30 days, secondary infections occurred in 13.3% of septic patients and 25.0% of non-septic patients (p=0.11). Although unadjusted frequencies were higher in non-septic patients, subsequent logistic regression showed no significant association between antibiotic-free days and secondary infection at 30 days. Furthermore, attempts to model in-hospital mortality were limited by perfect separation, precluding a reliable estimate of mortality risk based on antibiotic-free days.

Multi-drug resistant organisms (MDROs) were defined as bacterial cultures with reported resistance to more than one antibiotic on microbial cultures. Among microbial culture-positive secondary infections following sepsis, 66.6% (n = 8) of isolated organisms were MDROs within 30 days of infection, while 48% (n = 12) of organisms cultured between 30 days and one year after study enrollment were MDROs.

In contrast, among non-septic patients, 33.3% (n = 6) of organisms cultured within 30 days of enrollment were MDROs, whereas 60% (n = 6) of organisms isolated between 30 days and one year after enrollment were MDROs.

Neither 30-day mortality (21.1% vs. 13.9%, p=0.35) nor readmission rates among survivors (14.4% vs. 8.3%, p=0.35) differed significantly between groups. In keeping with the regression findings, antibiotic-free days were also not significantly linked to readmission risk at 30 days (p>0.05). The median hospital length of stay (LOS) was 9.0 days (IQR 5–14.3) for septic patients and 9.0 days (IQR 5.25–18.8) for non-septic patients (p=0.37). **Supplementary Fig 1** illustrates the overall LOS distribution for the entire cohort, demonstrating a non-normal distribution with most patients staying fewer than 15 days.

### Long-Term Outcomes

Kaplan-Meier survival curves (**Fig 2**) demonstrate that patients in the long-duration group (≥8 days of antibiotics, n=69) had similar one-year survival rates compared to those with short (4–7 days, n=36) or ultra-short (≤3 days, n=21) antibiotic courses (log-rank p>0.05).

Consistent with our logistic regression findings, longer antibiotic duration did not significantly affect one-year mortality. Multivariate modeling confirmed no difference in one-year mortality across these antibiotic-duration categories after adjusting for demographic and clinical covariates. By one year, all-cause mortality rates were likewise similar in septic (37.8%) and non-septic (36.6%) patients (p=0.86). Among one-year survivors, place of residence did not differ substantially between the two groups, although septic patients were numerically more likely to be discharged home (60.0% vs. 44.4%, p=0.11). Neither sepsis status nor antibiotic-free days significantly predicted one-year mortality (p>0.05), and neither sepsis status nor primary infection type emerged as significant predictors of secondary infections during long-term follow-up.

### Antibiotic Duration and Secondary Infections

**Fig 1** depicts the relationship between AD during the index hospitalization and the number of secondary infection episodes occurring within one year. Median AD increased with a higher number of subsequent infection episodes (Kruskal-Wallis p<0.05); however, post-hoc Dunn’s tests yielded mixed findings depending on the specific pairwise comparisons. Logistic regression adjusting for age, APACHE II score, gender, and glucocorticosteroid dose confirmed that longer antibiotic duration correlated with an increased hospital LOS (p<0.001) but showed no significant association with secondary infection at 30 days (p>0.05). Each additional day of antibiotics was associated with an estimated 0.37–0.39 extra days of hospitalization in univariate and multivariate analyses, respectively.

**Fig 1.**
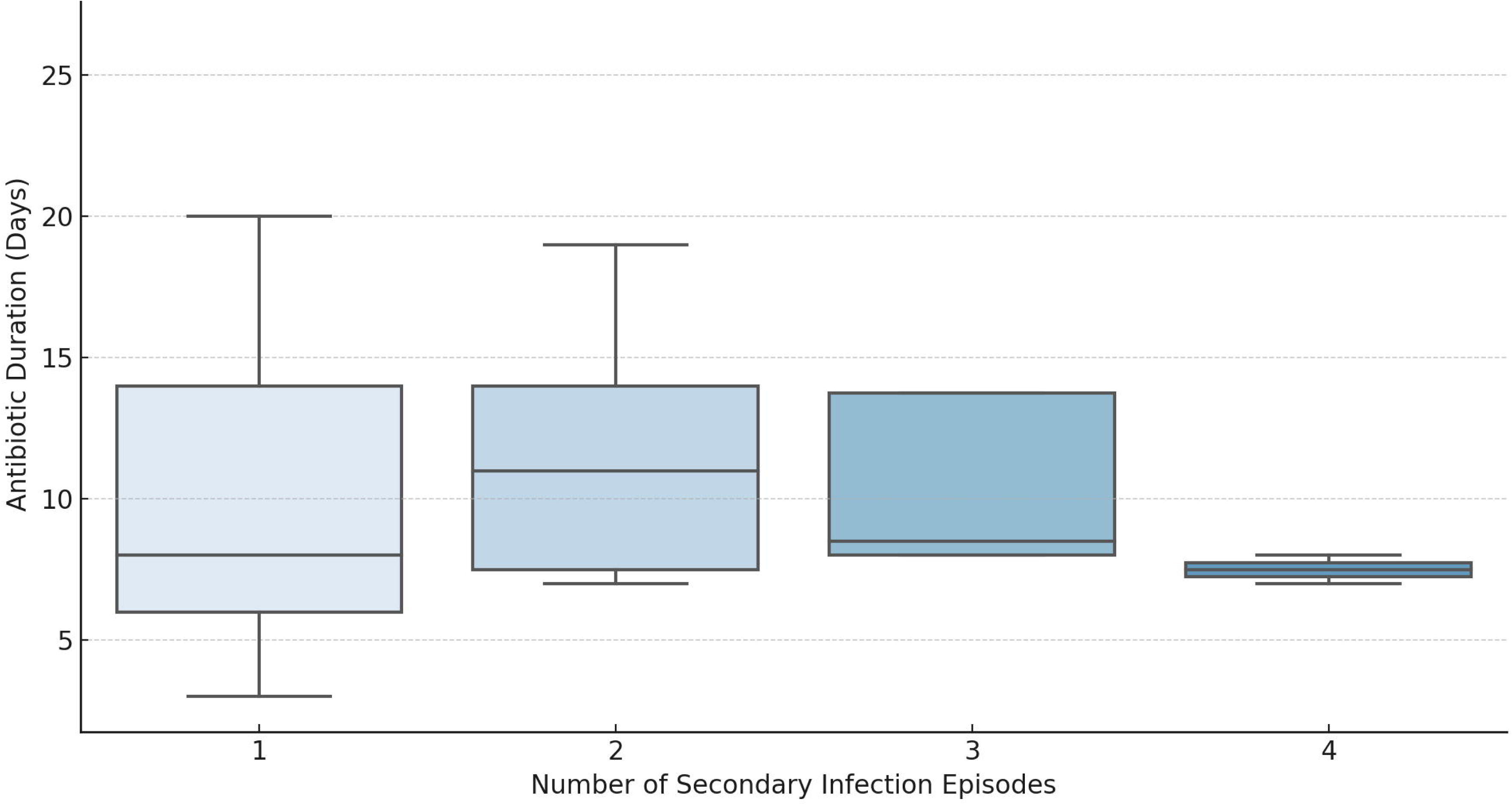
Relationship between the duration of antibiotic use during the index hospitalization and the number of secondary infection episodes within one year. Boxes display the interquartile range (IQR) with the median denoted by a horizontal line. Whiskers indicate 1.5 times the IQR, and outliers are represented by dots. Statistical significance was assessed using a Kruskal-Wallis test to evaluate differences in antibiotic duration across groups. Post-hoc pairwise comparisons were conducted using Dunn’s test with Bonferroni correction for multiple comparisons. A p-value <0.05 was considered statistically significant.

**Fig 2.**
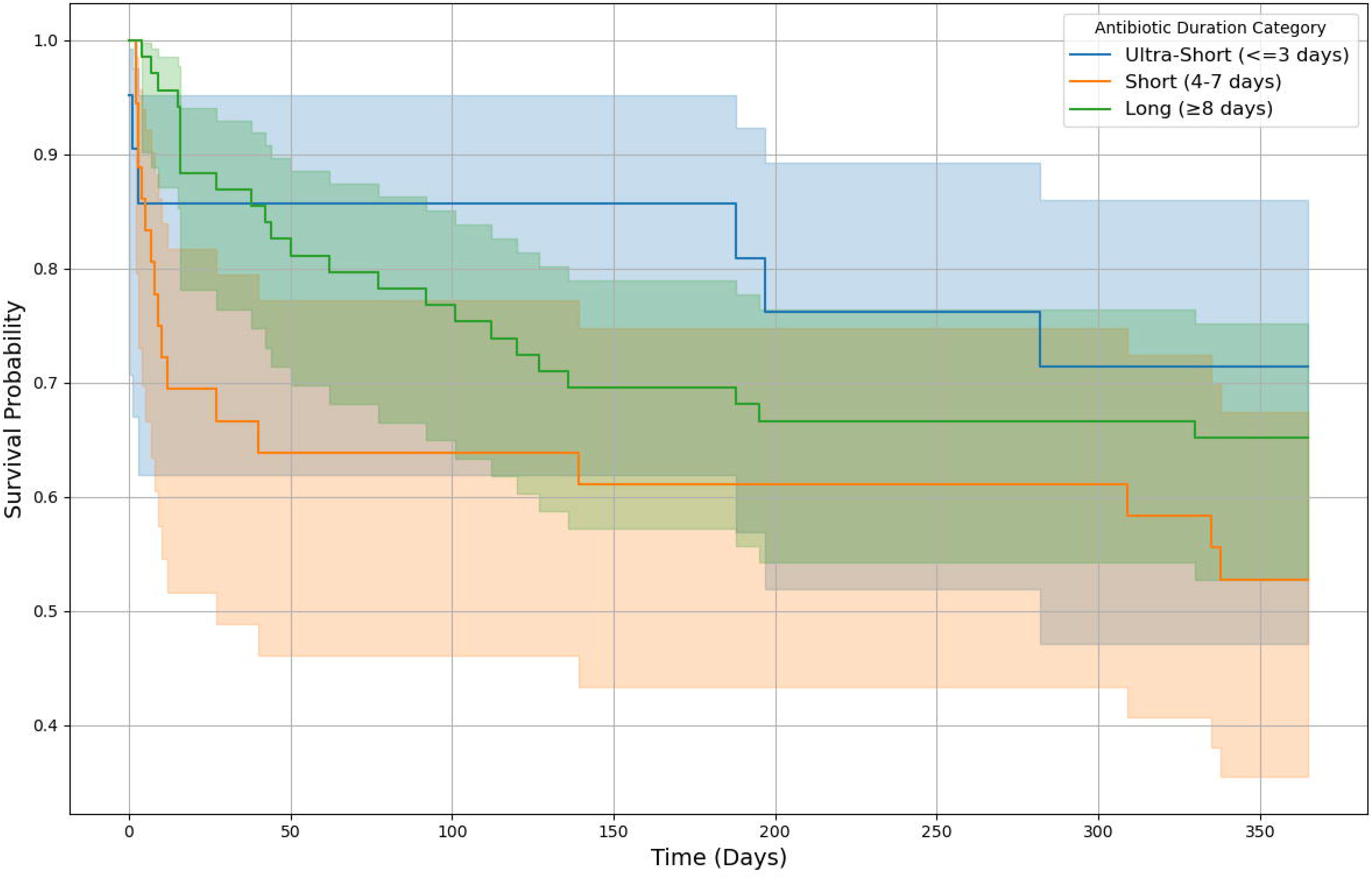
One-Year Survival by Antibiotic Duration During Index Hospitalization. This Kaplan-Meier survival curve illustrates the probability of one-year survival stratified by antibiotic duration (AD) categories during the index hospitalization. The shaded regions represent 95% confidence intervals. Patients who survived beyond one year were censored at 365 days. In the long-duration group (≥8 days), 69 patients were included, with 24 deaths at one year. In the short-duration group (4–7 days), 36 patients were included, of whom 17 died within one year. In the ultra-short group (≤3 days), 21 patients were included, with 6 deaths at one year. There was no significant difference between AD and mortality, after factoring age, APACHE II score, gender and glucocorticosteroid dose as co-variates.

Despite the absence of a significant association between AD and mortality, the original unadjusted findings suggested higher secondary bacteremia rates with fewer antibiotic-free days, but adjusted analyses did not confirm a statistically significant effect. **Supplemental Fig 2** further illustrates the inverse relationship between AFD and secondary infection risk among one-year survivors, stratified by primary infection source, though these findings should be interpreted with caution given the non-significant results of our multivariable models.

### Subgroup Analyses

In septic patients specifically, although unadjusted analyses indicated that fewer antibiotic-free days were associated with higher rates of secondary bacteremia (p=0.030), this relationship was no longer significant after adjusting for age, APACHE II score, and other covariates. Sepsis status was not an independent determinant of either short- or long-term mortality, nor did it alter the overall patterns in AD or one-year readmissions. Infection etiology (gram-positive vs. gram-negative vs. fungal vs. viral) did not influence AD or secondary infection patterns in either the septic or non-septic subgroup.

## Discussion

The findings from this study highlight how AD can shape clinical outcomes in critically ill patients, including those with sepsis. Although prolonged AD significantly increased hospital LOS, it did not affect mortality or readmission rates at either 30 days or one year, nor did it alter discharge disposition. While unadjusted analyses linked fewer antibiotic-free days to a higher incidence of secondary infections—particularly bacteremia—those associations lost significance after adjustment, suggesting that factors beyond antibiotic exposure alone may contribute to secondary morbidity. Collectively, these observations underscore the importance of prudent antibiotic management and illuminate the potential effects of extended therapy on patient-centered outcomes, including hospital stay and subsequent infectious complications.^14^

Often, antibiotic courses are initiated in the absence of positive microbial culture results, driven by clinical suspicion of infection. While this empiric approach is necessary in many cases, especially in high-risk and critically ill patients, the potential benefit must be balanced against the risk of antibiotic overuse. Even short-duration courses can disrupt both whole-body and individual organ microbiomes, leading to immune dysregulation and increased susceptibility to secondary infections. In contrast, our data indicate that reducing AD does not compromise mortality or risk of readmission, suggesting that efforts to abbreviate antibiotic courses may confer clinical benefits without diminishing short- or long-term survival.

An equally prominent finding concerns the role of AFD in mitigating secondary infections, including bacteremia. While unadjusted analyses suggested that increasing AFD was associated with reduced bacteremia over one year, these associations did not remain statistically significant after accounting for confounders. Nonetheless, this observation in unadjusted models aligns with the broader shift toward antimicrobial stewardship practices aiming to curb collateral damage from prolonged therapies. In this context, patients in our short (4–7 days) and ultra-short (≤3 days) AD groups did not exhibit higher mortality after adjusting for relevant confounders, reinforcing the notion that shorter, carefully targeted antibiotic regimens may balance infection control with reduced harm.

Although septic patients generally present with greater illness severity, the adverse effects of prolonged AD—particularly extended LOS—were evident in both sepsis and non-sepsis groups. The initially observed higher rate of bacteremia in patients with fewer AFD did not persist after multivariable adjustment, suggesting that other factors may drive secondary infection risk irrespective of sepsis status. Notably, primary infection type did not differentially influence trends in AD, AFD, or secondary infection outcomes. Rather, these results indicate that extended antibiotic courses can adversely affect critically ill patients regardless of underlying microbial etiology, potentially via antibiotic-induced immunoparalysis.

These data are consistent with prior trials evaluating abbreviated ADs. The BALANCE and SAFE trials have demonstrated that shorter courses often yield non-inferior outcomes for specific infections, such as bloodstream infections.^15–17^ Meta-analyses of community-acquired pneumonia^16,18–21^ and other infections^22–26^ further support that antibiotic regimens of seven days or fewer are safe and effective for many clinical scenarios. More targeted investigations focused on pathogens such as *Staphylococcus aureus*, vancomycin-resistant enterococci, and *Pseudomonas aeruginosa* corroborate that reduced durations do not necessarily worsen survival or readmission rates.^27–30^ The current study extends these observations by examining both confirmed and unknown microbial etiologies in critically ill populations, thereby highlighting that the adverse effects of prolonged AD on LOS and secondary infections persist even when the causative organism is unclear.

Despite the insights gained, several limitations must be acknowledged. As a retrospective, non-randomized study reliant on historical data, there is a possibility of measurement error and unmeasured confounding, despite adjustments for age, APACHE II score, and corticosteroid use. Additionally, the relatively small cohort may limit broader generalizability. Nonetheless, this study contributes value by describing long-term outcomes, which are infrequently examined in antibiotic literature. Furthermore, variability in infection source, severity, and comorbidities could influence prescribing patterns independently of illness severity. However, the consistent association of prolonged AD with increased LOS and secondary infections across multivariate and sensitivity analyses suggests a robust signal.

## Conclusion

These findings underscore the importance of thoughtful antibiotic stewardship in critically ill patients. Although the optimal threshold for “too long” or “too short” remains debated, minimizing antibiotic overexposure and maximizing AFD appear to be prudent strategies for reducing iatrogenic complications. Our results show that extended antibiotic duration prolongs hospital stays without improving mortality or readmission. Future prospective trials and de-escalation protocols are essential for clarifying the balance between ensuring adequate treatment and mitigating adverse effects of excessive antibiotic use. In parallel, additional research into immunoparalysis and host-microbe interactions may elucidate the underlying mechanisms of these observations. Ultimately, these data reinforce the value of robust stewardship programs focused on individualized antibiotic prescribing to optimize outcomes for critically ill patients and limit the unintended consequences of antibiotic overuse.

## Methods

### Study Design and Ethical Considerations

This study was conducted as a retrospective cohort analysis approved by the Institutional Review Board (IRB No. 15328, approved July 30, 2020) at Penn State College of Medicine. All procedures adhered to the Declaration of Helsinki and local institutional guidelines. Patients admitted between April 2023 and July 2024 who were flagged by a Modified Early Warning Score (MEWS)-based algorithm were screened for possible sepsis. Two investigators independently reviewed flagged patients to minimize selection bias and confirmed eligibility based on clinical data. Written informed consent was obtained from patients with decision-making capacity or from legally authorized representatives before enrollment.

### Study Population and Screening

Patients were included if they were 18 years of age or older, identified within 48 hours of critical illness onset, and met Sepsis-3 criteria, characterized by a two-point increase in Sequential Organ Failure Assessment (SOFA) score in the context of suspected or confirmed infection.^31,32^ Critical illness was defined by the need for continuous intravenous vasopressors or noninvasive or invasive respiratory support. Patients were excluded if they had comfort measures only, were admitted under hospice care, were pregnant, were part of a protected population lacking IRB permission, or had incomplete records preventing calculation of severity scores.

### Data Collection and Management

Electronic health records (EHR) provided demographic data, clinical information, medication records (including AD), vital signs, laboratory values, and outcomes. Structured post-discharge interviews were conducted to resolve ambiguities or missing information and to validate events such as hospital readmissions or additional antibiotic use after discharge. Data were extracted from secure EHR queries and subsequently loaded and analyzed using Python v.3.9.16. Missing data were imputed where feasible, and any records with critical fields missing were flagged for potential exclusion. Variables of interest included AD, calculated as the total number of days on antibiotics during the index admission, and AFD, determined by subtracting AD from the length of observation (up to hospital discharge or 30 days, whichever occurred first). Covariates entered into multivariate analyses included age, gender, body mass index (BMI), Charlson Comorbidity Index,^33^ APACHE II score,^34^ and glucocorticosteroid use or dose. Severity metrics such as SOFA and APACHE II were generated using standardized scoring algorithms.

### Outcome Definitions

The primary outcome was secondary infection, defined as a new episode requiring at least three consecutive days of antibiotic therapy, initiated after the completion of the primary course and within one year of the index sepsis admission. Secondary outcomes included one-year bacteremia (confirmed via blood cultures), other positive cultures (e.g., urine, tissue, respiratory), one-year hospital readmissions (infection-related and non-infection-related), one-year all-cause mortality, index hospital LOS, and discharge disposition (home, rehabilitation, skilled nursing facility, or other). These outcomes were monitored through thorough review of EHR records, supplemented by structured phone interviews conducted at 30 days, 3 months, 6 months, and 12 months following study enrollment.

### Statistical Analysis

Descriptive statistics summarized categorical variables as counts and proportions, with comparisons made via Pearson’s Chi-square or Fisher’s exact test. Continuous variables were presented as means with standard deviation or medians with interquartile ranges, depending on their distribution; these were compared using unpaired t-tests or nonparametric alternatives.

Multivariate logistic regression models were used to explore the effects of AD and AFD on the primary and secondary outcomes, with adjustments for potential confounders such as age, APACHE II score, Charlson Comorbidity Index, and glucocorticosteroid use. The script included variable standardization (mean centered and scaled to unit variance) if required, with model diagnostics such as pseudo-R², Hosmer-Lemeshow tests, and confusion matrices generated to evaluate model fit. Sensitivity analyses examined septic versus non-septic subgroups and accounted for different definitions of secondary infection (for instance, requiring microbiological confirmation). Additional tests addressed interactions between antibiotic exposure and corticosteroid use. Multiple comparisons were accounted for by adjusting p-values, and missing data were handled with imputation or case-wise exclusions when critical fields were incomplete. Results were reported as odds ratios (OR) with 95% confidence intervals (CI).

### Follow-up and Quality Control

Patients and, if necessary, their families were contacted at predefined intervals to document changes in health status, readmissions, and antibiotic use. If telephone follow-up was unsuccessful, the EHR was rechecked to ensure that relevant information was captured. A subset of patient data was independently entered and coded by two researchers to evaluate inter-rater reliability via Cohen’s kappa, ensuring that variables such as infection type and sepsis classification were consistently assigned. Python scripts were version-controlled to maintain a transparent audit trail of data cleaning, variable construction, and statistical analyses. This systematic approach minimized bias and maximized reproducibility, forming the foundation for subsequent interpretation of the associations between AD, AFD, and clinical outcomes.

## Data Availability

All data produced in the present study are available upon reasonable request to the authors.

