## Supplementary Material for "Association Between In-Hospital Antibiotic Use and Long-Term Outcomes in Critically Ill Patients"

**Supplementary Materials**

**Supplementary Table 1.** Microbial etiology and infectious source within the study cohort

| **Name of Organism cultured** | **Blood** | **Urine** | **Respiratory** | **Abdomen** | **Soft tissue/skin** |
| --- | --- | --- | --- | --- | --- |
| *Actinomyces odontolyticus* |  |  |  | 1 |  |
| *Aerococcus urinae* |  | 1 |  |  |  |
| *Anaerobic gram-positive rods (unspeciated)* |  |  |  | 1 |  |
| *Bacteroides fragilis* | 1 |  |  |  |  |
| *Bacteroides pyogenes* |  |  |  | 1 |  |
| Bacteroides thetaiotaomicron |  |  |  | 1 |  |
| Beta streptococcus, group B | 4 |  | 1 |  |  |
| Beta streptococcus, group G | 1 |  |  |  |  |
| Beta streptococcus, group A | 3 |  |  |  | 1 |
| *Bifidobacterium* |  |  |  | 1 |  |
| *Candida albicans* |  |  |  | 1 |  |
| *Candida (unspeciated)* |  |  | 1 |  |  |
| *Candida glabrata* | 3 | 1 | 1 | 1 |  |
| *Candida lusitaniae* |  |  | 1 | 1 |  |
| *Clostridium clostridiforme* |  |  |  | 1 |  |
| *Clostridium difficile* |  |  |  | 1 |  |
| *Clostridium perfringens* |  |  |  | 2 |  |
| *Corynebacterium striatum* | 1 |  |  |  |  |
| SARS-CoV-2 |  |  | 5 |  |  |
| *Enterobacter cloacae* | 1 |  | 1 | 1 |  |
| *Enterococcus* (unspeciated) | 1 |  |  |  |  |
| *Enterococcus faecalis* |  | 2 |  | 1 |  |
| *Enterococcus faecium* |  | 2 |  | 1 |  |
| *Escherichia coli* | 8 | 6 | 2 | 5 | 1 |
| *Fusobacterium nucleatum* | 1 |  |  |  |  |
| *Gemella morbillorum* | 1 |  |  |  |  |
| Gram-negative rods (unspeciated) |  |  | 2 |  |  |
| *Haemophilus influenza* |  |  | 2 |  |  |
| Influenza A |  |  | 2 |  |  |
| *Klebsiella oxytoca* | 2 | 1 | 1 |  |  |
| *Klebsiella pneumoniae* | 4 | 4 |  | 1 |  |
| *Staphylococcus aureus* | 4 |  | 7 |  | 5 |
| *Morganella morganii* | 1 |  |  | 1 | 1 |
| *Prevotella intermedia* | 1 |  |  |  | 1 |
| *Proteus mirabilis* | 5 | 6 |  |  |  |
| *Proteus vulgaris* | 1 |  |  |  |  |
| *Pseudomonas aeruginosa* |  | 1 | 2 |  |  |
| Rhinovirus/enterovirus |  |  | 1 |  |  |
| *Rhodotorula* |  |  | 1 |  |  |
| *Serratia marcescens* |  | 1 |  |  |  |
| *Staphylococcus epidermidis* (two positive blood cultures) | 2 |  |  |  |  |
| *Staphylococcus lugdenensis* | 2 |  | 1 |  |  |
| *Streptococcus anginosus* | 1 |  |  |  | 1 |
| *Streptococcus mutans* | 1 |  |  |  |  |
| *Streptococcus viridans* |  |  | 1 |  |  |

* If multiple microbes were isolated from a patient's microbial cultures, each microbe is listed individually.

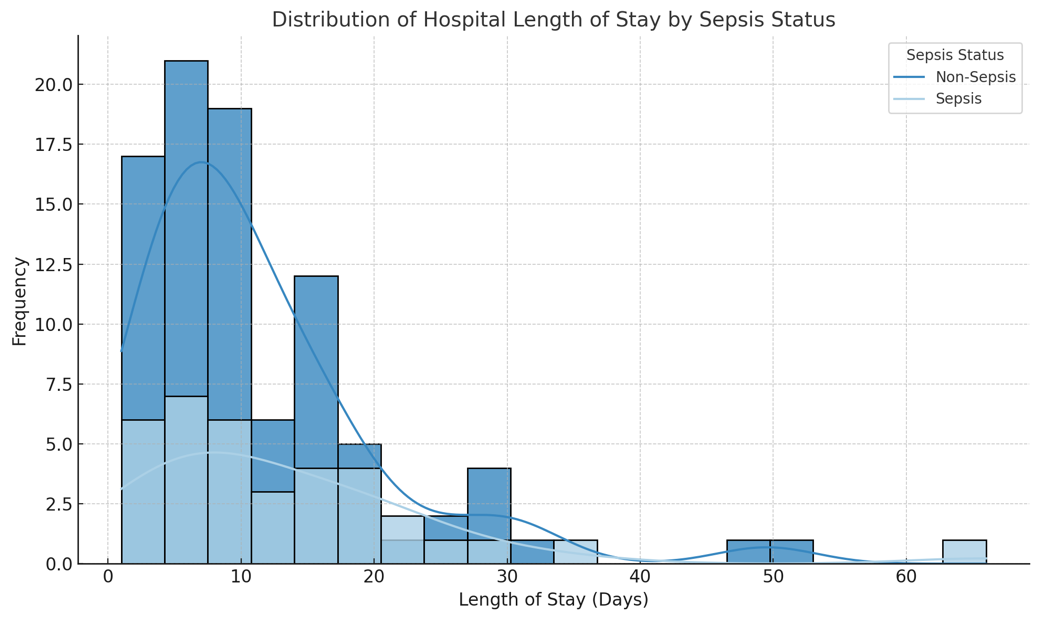

**Supplementary Figure 1.** **Distribution of index hospital length of stay (LOS) among the study cohort.** The x-axis represents LOS in days, while the y-axis shows the frequency of patients within each bin. Bins were set to 5-day intervals to capture variation while maintaining interpretability. Descriptive statistics include mean LOS (11.9 days), median (9 days), and standard deviation (10.1 days). Statistical analysis of LOS was performed using the Shapiro-Wilk test to assess normality. Based on non-normal distribution, the Mann-Whitney U test was employed to compare LOS between groups (e.g., sepsis vs. non-sepsis patients). Data suggest considerable heterogeneity in LOS, with most patients staying fewer than 15 days.

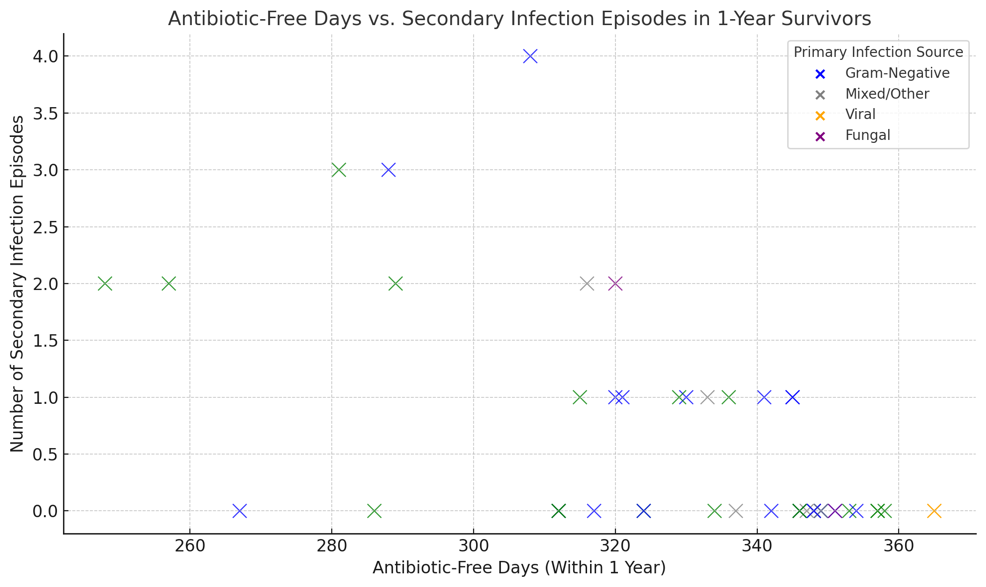

**Supplementary Figure 2. Relationship between antibiotic-free days and secondary infection, in 1-year survivors**, stratified by primary infection source. Each dot represents one patient; n=49.
